# Predictive modelling of COVID-19 New Confirmed Cases in Algeria using Artificial Neural Network

**DOI:** 10.1101/2021.03.29.21254532

**Authors:** Messaoud Djeddou, Ibrahim A. Hameed, Aouatef Hellal, Abolfazel Nejatian

**Affiliations:** Research Laboratory in Subterranean and Surface Hydraulics (LARHYSS), Faculty of Sciences and Technology, Mohamed Khider University of Biskra, Algeria. PO box 145 RP, 07000 Biskra, Algeria; Department of ICT and Natural, Sciences, Faculty of Information Technology and Electrical Engineering, Norwegian University of Science and Technology (NTNU), Postboks 1517, 6025 Ålesund, Norway; Department of Electrical Engineering, Florida Polytechnic University, FL, USA

**Keywords:** COVID-19, Predictive modeling, Artificial neural networks, Multi-layer perceptron, Radial basis function

## Abstract

This study investigates the potential of a simple artificial neural network for the prediction of COVID-19 New Confirmed Cases in Algeria (CNCC).

Four different ANN models were built (GRNN, RBFNN, ELM, and MLP). The performance of the predictive models is evaluated based on four numerical parameters, namely root mean squared error (RMSE), mean absolute error (MAE), Nash-Sutcliffe efficiency (NSE), and Pearson correlation coefficient (R). Taylor diagram was also used to examine the similarities and differences between the observed and predicted values obtained from the proposed models.

The results showed the potential of the multi-layer perceptron neural network (MLPNN) which exhibited a high level of accuracy in comparison to the other models.

## I. Introduction

Bacteria, parasites and viruses, can cause serious diseases for human’s health but the most serious diseases are usually caused by pathogenic viruses and among these viruses, coronavirus are the large family of pathogenic viruses.

Infection with these types of viruses can cause respiratory, liver, gastrointestinal and neurological diseases. They are distributed among humans, birds, cattle, mice, bats and other wildlife [1–2].

In December 2019, the World Health Organization (WHO) received notifications from Chinese authorities for many respiratory illnesses linked to people who visited a local market for seafood and wildlife in Wuhan city, Hubei Province, China [3]. Virological investigation suggests that the causative agent of this pneumonia is a new coronavirus (COVID-19) [4].

The city of Wuhan has become the main spread focus of a new coronavirus called COVID-19 (previously it was called 2019-nCoV), this new coronavirus has made hundreds of thousands of victims around the world, and become the global pandemic threatening human health.

Different researchers employed statistical and mathematical modeling approaches to predict the spreading of the outbreak in different countries.

Dehesh et al. [6] proposed an Auto Regressive Integrated Moving Average (ARIMA) model to predict the trend of confirmed cases in different countries using data collected from Johns Hopkins University official website.

Furthermore,

Zhao et al. [7] proposed a mathematical model to estimate the real number of COVID-19 cases in the first half of January 2020. They concluded that the number of unreported cases resulted in 469 cases from 1 to 15 January 2020 determined by the maximum likelihood estimation.

Nishiura et al. [8, 10] proposed an estimation model for the infection rate of COVID-19 in Wuhan, China using the data of 565 Japanese citizens who had been evacuated from Wuhan from 29 to 31 January 2020.

Machine learning and artificial intelligence have become very important tools in real applications especially with the era of big data.

Al-qaness et al. [9] proposed an improved adaptive neuro-fuzzy inference system (ANFIS) to forecast the COVID-19 confirmed cases of the upcoming ten days. The outcomes showed good performances.

Genetic evolutionary programming has been applied to forecast the spread of COVID-19 outbreak in India. The study show the capabilities of the proposed model to forecasting both total cases and death cases with highly accuracy [10]. A NARANN model was applied to forecast the total COVID-19 cases in Egypt. The model has a better accuracy than that of conventional ARIMA model [11].

Artificial neural network-based curve fitting techniques was used for prediction and forecasting of the Covid-19 number of rising cases and death cases in India, USA, France, and UK. The results have shown that ANN can efficiently forecast the future cases of COVID 19 outbreak of any country [12].

A multi-layer perceptron neural network (MLPNN) model was proposed to estimate and forecast the number of confirmed and recovered cases of COVID-19 in the upcoming days until September 17, 2020 in KSA [13].

An extreme learning machine (ELM) was used to predict the COVID-19 new cases in Algeria. The results showed that the proposed ELM model achieved an adequate level of prediction accuracy with smallest errors [14].

In this study, four artificial neural network models namely (radial basis function neural networks “RBFNN”, generalized regression neural networks (GRNN), Extreme learning machine (ELM), and multi-layer perceptron neural network (MLPNN) are proposed for predictive modeling of COVIS-19 new confirmed cases (CNCC) in Algeria.

## II. MATERIAL AND METHODS

### A. Case study

The pandemic Covid-19 in Algeria spreads from February 25, 2020, when an Italian citizen was tested positive for SARS-CoV-2, and then from March 1, an outbreak of contagion is formed in the department of Blida [15]. Sixteen members of the same family were infected with the coronavirus at a wedding party following contact with Algerian nationals in France [16]. The Blida department becomes the epicenter of the coronavirus epidemic in Algeria [17].

Confirmed Covid-19 cases was then detected, there were 2769 deaths and 100195 confirmed cases in Algeria as of January 3rd, 2021.

### B. Multi-layer perceptron neural network (MLPNN)

Artificial Neural Networks (ANNs) present a very flexible, robust and general method of modeling data. An ANN can be trained on experimental data to model a nonlinear mapping realized by some system. One of the advantages of an ANN model, over traditional nonlinear regression approaches, is that it is not necessary to first find a suitable parametric form of the regression model for the problem at hand. A one hidden layer multilayer perceptron can act as a universal function approximator [19]. A schematic diagram of the MLPNN architecture is shown in figure 2.

**Fig. 1.**
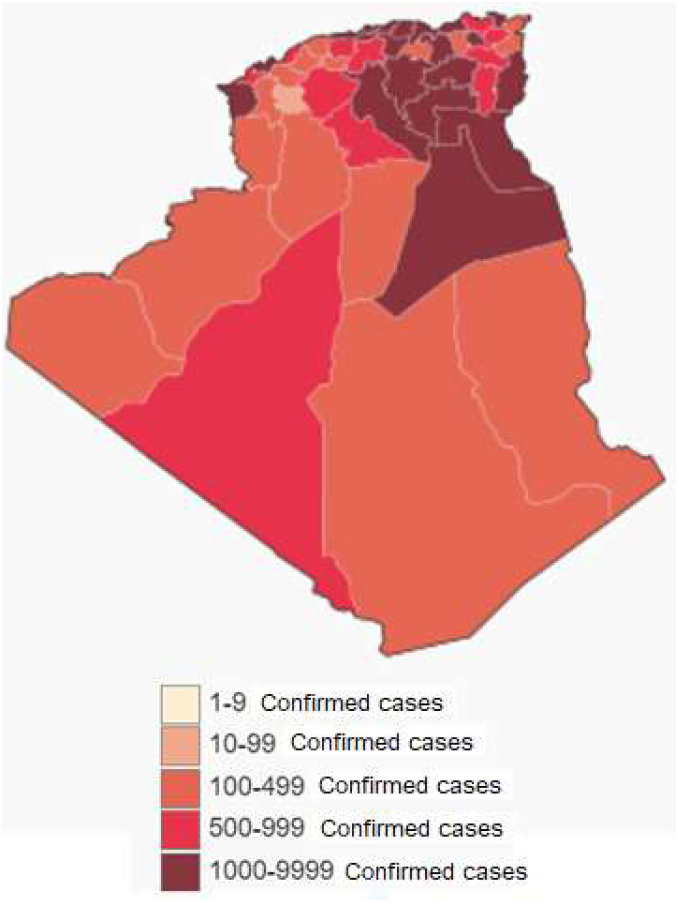
Map of the Covid-19 pandemic in Algeria [18].

**Fig. 2.**
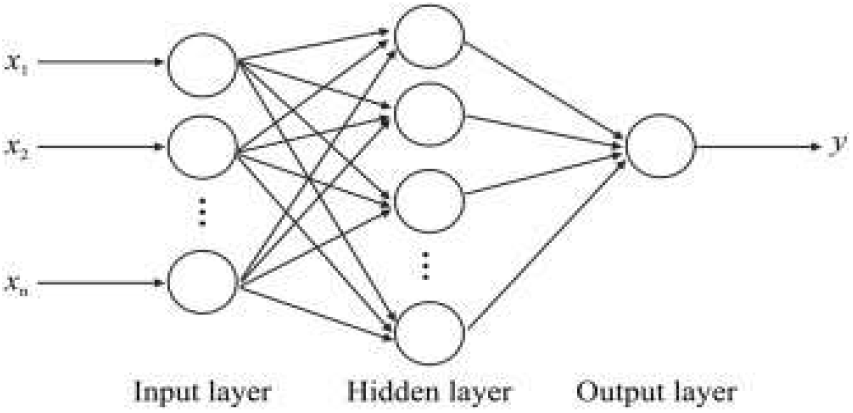
STRUCTURE OF A MULTI-LAYER PERCEPTRON NEURAL NETWORK (MLPNN)

ANNs may be defined as structures comprised of densely inter-connected adaptive simple processing elements (called artificial neurons or nodes) that are capable of performing massively parallel computations for data processing and knowledge representation [20-21].

### C. Radial basis function neural network (RBFNN)

Radial basis function neural networks (RBFNN) are widely used for predictive modeling in engineering fields [22-23]. RBFNN as show in figure 3, is represented as a three-layered architecture. First layer is the input layer that receives the inputs. The middle layer is known as the hidden layer, which contains a non-linear RBF activation function. The third one is the output layer that makes the prediction. The formulation of a RBFNN output is as follows:

**Fig. 3.**
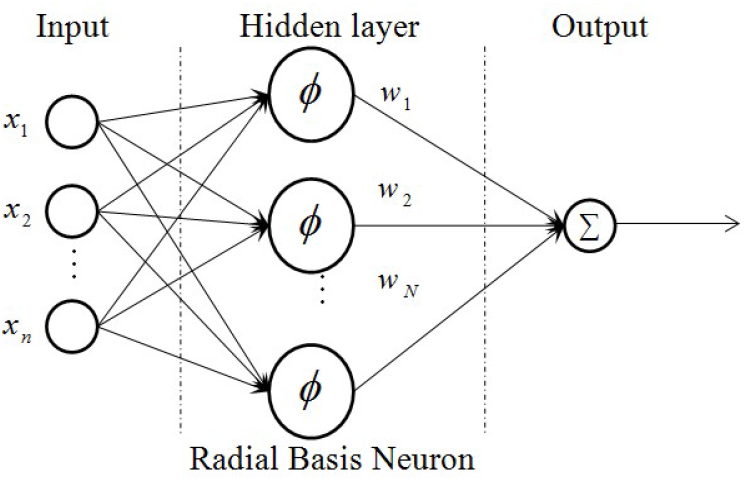
STRUCTURE OF A RADIAL BASIS FUNCTION NEURAL NETWORK (RBFNN)

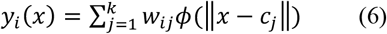

Where: *x* is input vector, *y*_*i*_ is the network’s *i*^th^ output, *k* is the number of neurons in the hidden layer, *c*_*j*_ denotes the center of the *j*^th^ hidden neuron, *w*_*ij*_ represents the weight of the link from the *j*^th^ neuron in the hidden layer to the *i*^th^ neuron in the output layer, and ‖. ‖ is the Euclidian norm; *ϕ* is the radial basis function which is used in the neurons of hidden layer.

This parameter is a multidimensional radial basis function describing the difference between an input vector and a pre-defined center vector. The most common radial basis functions used is the Gaussian Function which is defined as follows:

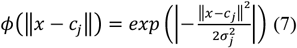

Where σj is the width of the jth hidden neuron, Finding the centers, widths, and the weights connecting hidden neurons to the output is the key for constructing and training the RBFNN. Both the dimensionality and the distribution of the input patterns affect the number of the hidden neurons. If the dimensionality is reduced, the number of hidden neurons will also be decreased [24].

Among the advantages of the RBFNN, their simple and compact architecture, which means there are fewer parameters that need to be optimized. The RBFN has good tolerance to the noise in the dataset and a high learning efficiency [25-26].

### D. Generalized Regression Neural Network (GRNN)

The Generalized Regression Neural Networks (GRNN) was firstly proposed by Specht 1991 [27]. A GRNN is a variation of the radial basis neural networks, which is based on kernel regression networks [28-29]. One of the advantages of this network is its consistency; the estimation error approaches zero, with only slight restrictions on the function as the size of the training set becomes large.

More details for the GRNN model are given in [27] and its schematic diagram is shown in figure 4.

**Fig. 4.**
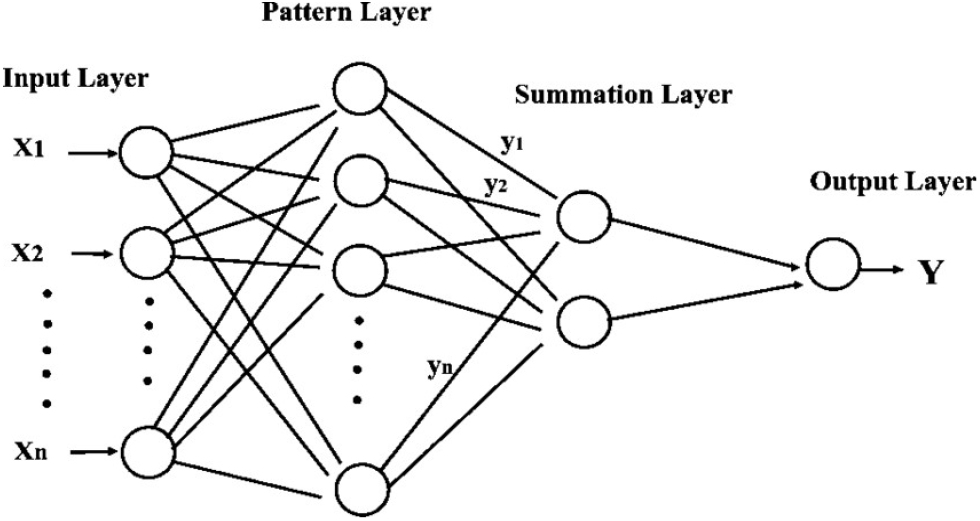
STRUCTURE OF A GENERALIZED REGRESSION NEURAL NETWORK (GRNN).

GRNN does not require an iterative training procedure as back propagation networks. It approximates any arbitrary function between input and output vectors, drawing the function estimate directly from the training data. In addition, it is consistent that as the training set size becomes large, the estimation error approaches zero, with only mild restrictions on the function [30].

A hidden neuron layer is created to hold the input vector. The weight between the newly created hidden neuron and the output neuron is assigned the target value. A GRNN is based on the following formula:

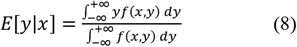

Where: *y* is the output of the estimator; *x* is the estimator input vector; *E*[*y*|*x*] is the expected value of output, given the input vector *x*, and *f(x,y)* is the joint probability density function (*pdf*) of *x* and *y*. The function value is estimated optimally as follows:

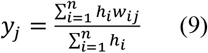

Where: *w*_*ij*_ is the target output corresponding to input training vector *x*_*i*_ and output *j*;

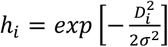: is the output of a hidden layer neuron;

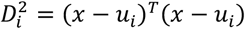: is the squared distance between the input vector *x*_*i*_ and the training vector *u*_*i*_; σ: a constant controlling the size of the perceptive region.

The major difference between GRNN and RBF neural networks is the method that the weights *w*_*ij*_, are determined. Instead of training weights, the GRNN assigns the target value directly to the weights *w*_*ij*_, from the training set associated with input training vector and a component of its corresponding output vector.

### E. Extreme learning machine (ELM)

The basic theory of ELM can be given as follows: For M arbitrary distinct inputs (xi,yi) with xi ∈ ℝd and yi ∈ ℝ; a standard SLFN with N hidden nodes and activation function f can be modelled as the following:

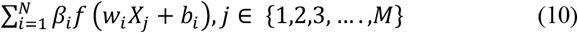

where wi are the input weights to the ith neuron in the hidden layer, bi the biases and bi are the output weights.

In the case where the SLFN would perfectly approximate the data, the relation is

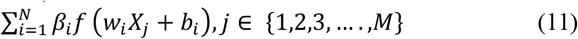

which can compactly be written as,

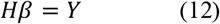

Where H the hidden layer output matrix is defined as,

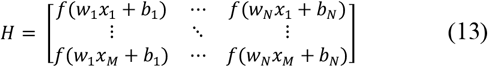

Where:

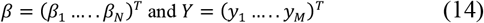

Considering the randomly initialized first layer of the ELM and the training inputs, the hidden layer output matrix H can be computed. Given H and the target outputs, output weight b can be solved by finding the least square solution to the linear system defined by Eq. (12). This solution is given by *b* = *H*^*ℳ℘*^*Y*, where *H*^*ℳ℘*^ is the Moore–Penrose generalized inverse of the matrix H. More details on the ELM algorithm can be found in the original paper [31-32].

### F. Performance Evaluation

The root mean square error (RMSE), mean absolute error (MAE), the Pearson correlation coefficient (R) and the Nash– Sutcliffe efficiency (NSE), which were applied in this study.

## III. RESULTS AND DISCUSSION

Four different models were developed for modeling the COVID-19 New Confirmed Cases (CNCC) in Algeria, as shown in Table 2. The models was developed using simple ANN (GRNN, RBFNN, ELM and MLPNN).

**TABLE I.** Descriptive statistics for the parameters used for New COVID-19 Cases Modelling Table Type Styles

| Parameters | Min. | Max. | Mean | Std. dev. |
| --- | --- | --- | --- | --- |
| **Inputs** |  |  |  |  |
| Cumulative confirmed Cases (*CCC*) | 3 | 100195 | 35003.3539 | 30462.3540 |
| Total Cumulative Deaths (*TCD*) | 0 | 2769 | 1244.5681 | 813.8320 |
| Daily Death Cases (*DDC*) | 0 | 30 | 8.97727 | 4.9716 |
| Daily Recovered Cases (*DRC*) | 0 | 649 | 219.51623 | 169.3326 |
| Cumulative Recovered Cases (*CRC*) | 0 | 67611 | 23635.9772 | 20533.3595 |
| Index Day (*ID*) | 1 | 7 | 4 | 2 |
| **Output** |  |  |  |  |
| COVID-19 New Confirmed Cases (*NCC*) | 0 | 1133 | 325.0811 | 261.3646 |

**TABLE II.**
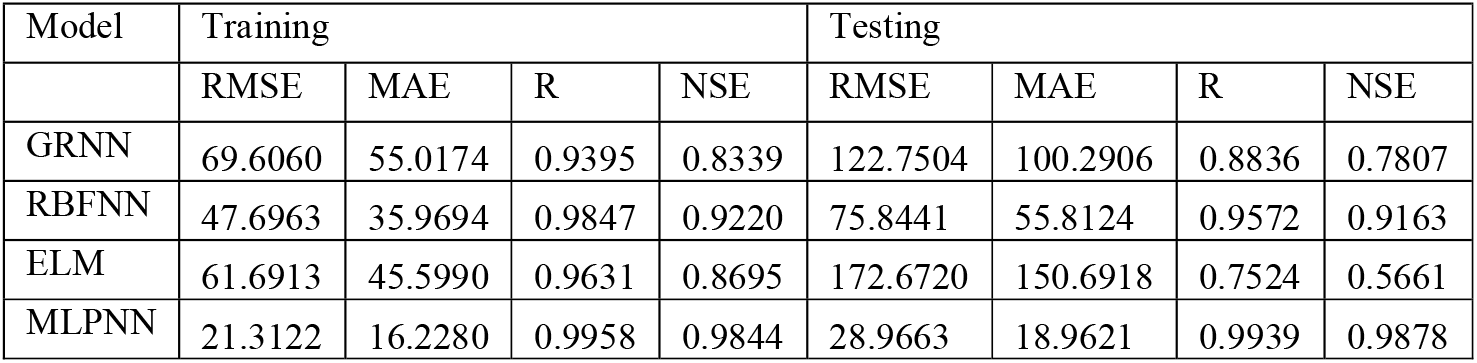
PERFORMANCE RESULTS OF GRNN, RBFNN, ELM, AND MLPNN MODELS.

The identification of a maximum number of hidden neurons, iterations, transfer function and better structure is a very important task to design the different ANN models in order to develop the best model. The development of all the models was performed under MATLAB 2019b environment.

The obtained results of the four models using GRNN, RBFNN, ELM and MLPNN models for the CNCC prediction are presented in Table 2.

From the overall comparison, it can be observed that all the model combinations demonstrate acceptable performance in the modeling horizon; this can be proved by considering the NSE values that are greater than 0.80 in both training and testing, except for ELM and GRNN models in testing phase.

Table 2 shows that the best model produces the highest prediction accuracy was MLPNN model in terms of RMSE, MAE, R and NSE for CNCC prediction. Among the models, MLPNN with, RMSE (21.3122), MAE (16.2280), R (0.9958) and NSE (0.9844) values in the training phase, had the highest level of accuracy in comparison to GRNN, RBFNN and ELM. The fitting between the observed and predicted CNCC is shown in figure 5. It can be clearly seen that the overall performance of MLPNN combinations is superior to GRNN, RBFNN, and ELM models combinations.

**Fig. 5.**
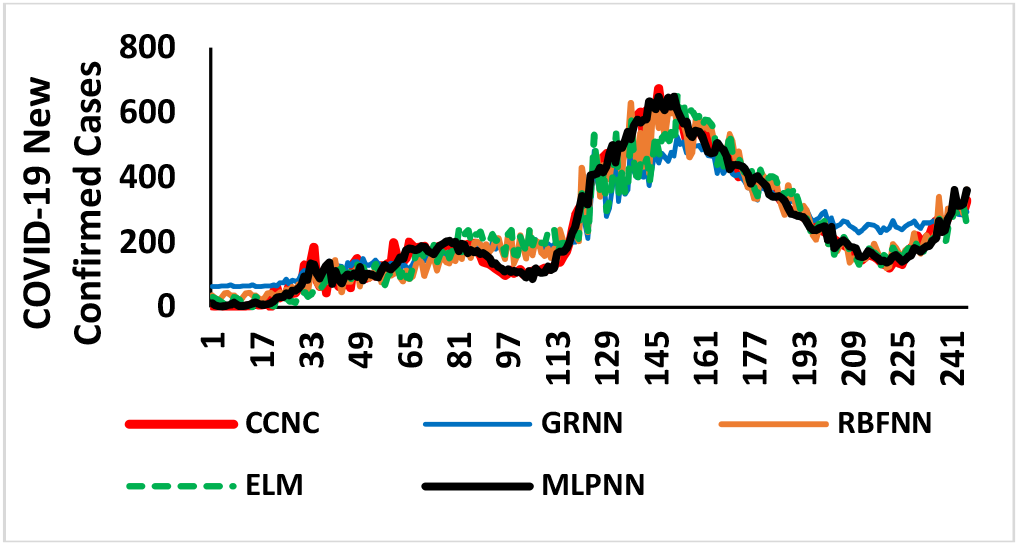
TRENDS OF PREDICTED CNCC PLOTS FOR THE GRNN, RBFNN, ELM AND MLPNN MODELS IN TRAINING PHASE.

Also, it is predicted values trends demonstrating better agreements with the observed values than other three models. In terms of overall percentage accuracy in training phase, MLPNN outperformed and increased the predictive performance up to 18.46%, 6.76% and 13.21% with regard to the GRNN, RBFNN and ELM models, respectively.

In testing phase, the MLP model outperforms GRNN, RBFNN and ELM by a relative small increase of the errors RMSE (28.9663), MAE (18.9621) and increase of performance parameters NSE (0.9878), respectively. The fitting between the observed and predicted CNCC values is shown in figure 6. The behavior of all models was influenced by the effect that highest values of CNCC are in testing phase.

**Fig. 6.**
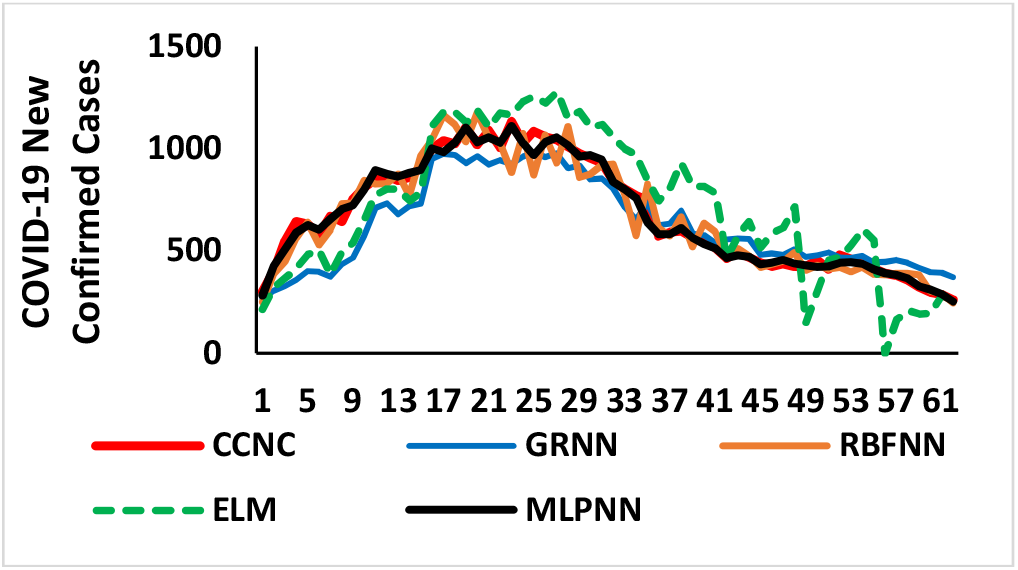
TRENDS OF PREDICTED CNCC PLOTS FOR THE GRNN, RBFNN, ELM, AND MLPNN MODELS IN TESTING PHASE.

The MLPNN model show superior generalization capabilities relative to GRNN, RBFNN, and ELM models.

It can be noted from the figure 7.d that predicted CNCC values using MLPNN are quite close to the observed CNCC values, as their R-square (R^2^) value is very close to unity.

**Fig. 7.**
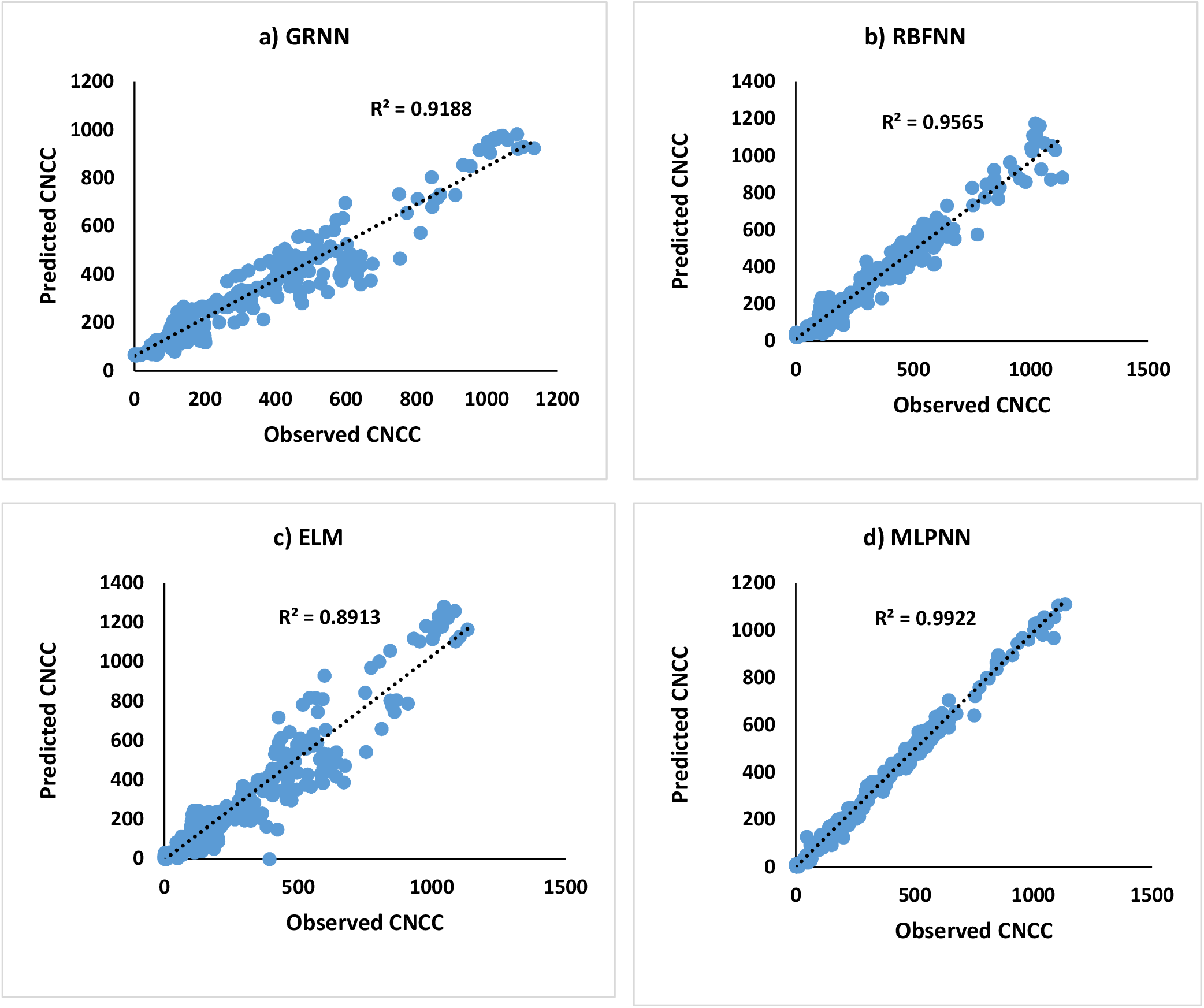
OBSERVED CNCC VS PREDICTED CNCC WITH GRNN, RBFNN, ELM AND MLPNN MODELS.

In order to capture the detail of the performance of the proposed predictive models using different ANN configuration (RBFNN, GRNN, ELM, and MLPNN), a two-dimensional method that exhibits how the proposed models matches the observed and corresponding predicted values of CNCC. Taylor diagram is constructed to visualize the information in figure 8.

**Fig. 8.**
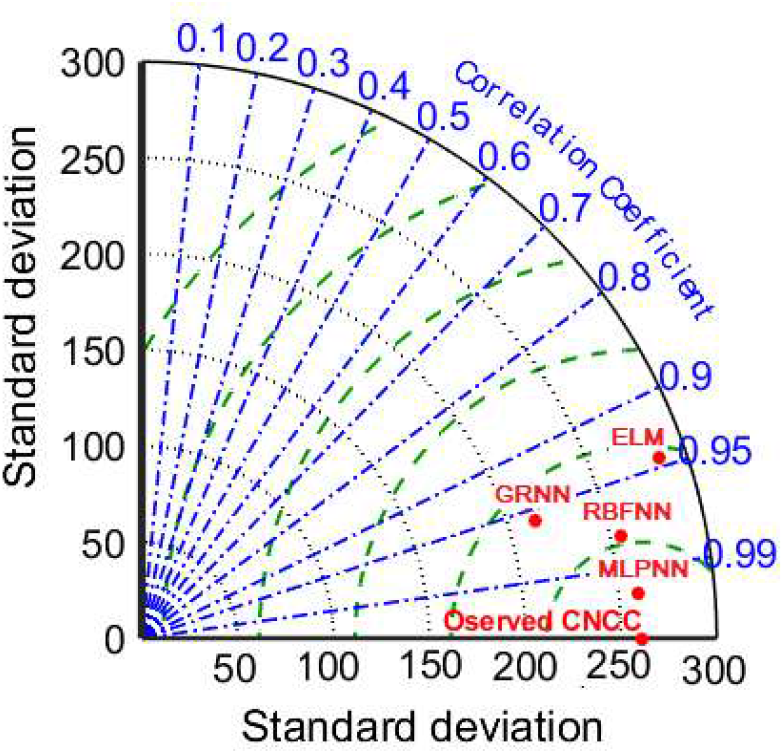
TAYLOR DIAGRAM DEPICTING THE BEST PERFORMANCE OF PREDICTIVE MODELING OF CNCC USING GRNN, RBFNN, ELM, AND MLPNN MODELS.

The Taylor diagram is also the most widely recommended diagram for accuracy comparison due to the advantageous nature of combining and quantifying multiple statistical performance metrics by comparing the similarity between the observed and predicted values in one diagram.

However, in accordance with the visualized graphical interpretation, the MLPNN model was closer to the target observed values in comparison with the other models.

Lastly, a comparative study is presented in figure 9, in terms of CNCC predicted values using the two best models in terms of performances (RBFNN and MLPNN).

**Fig. 9.**
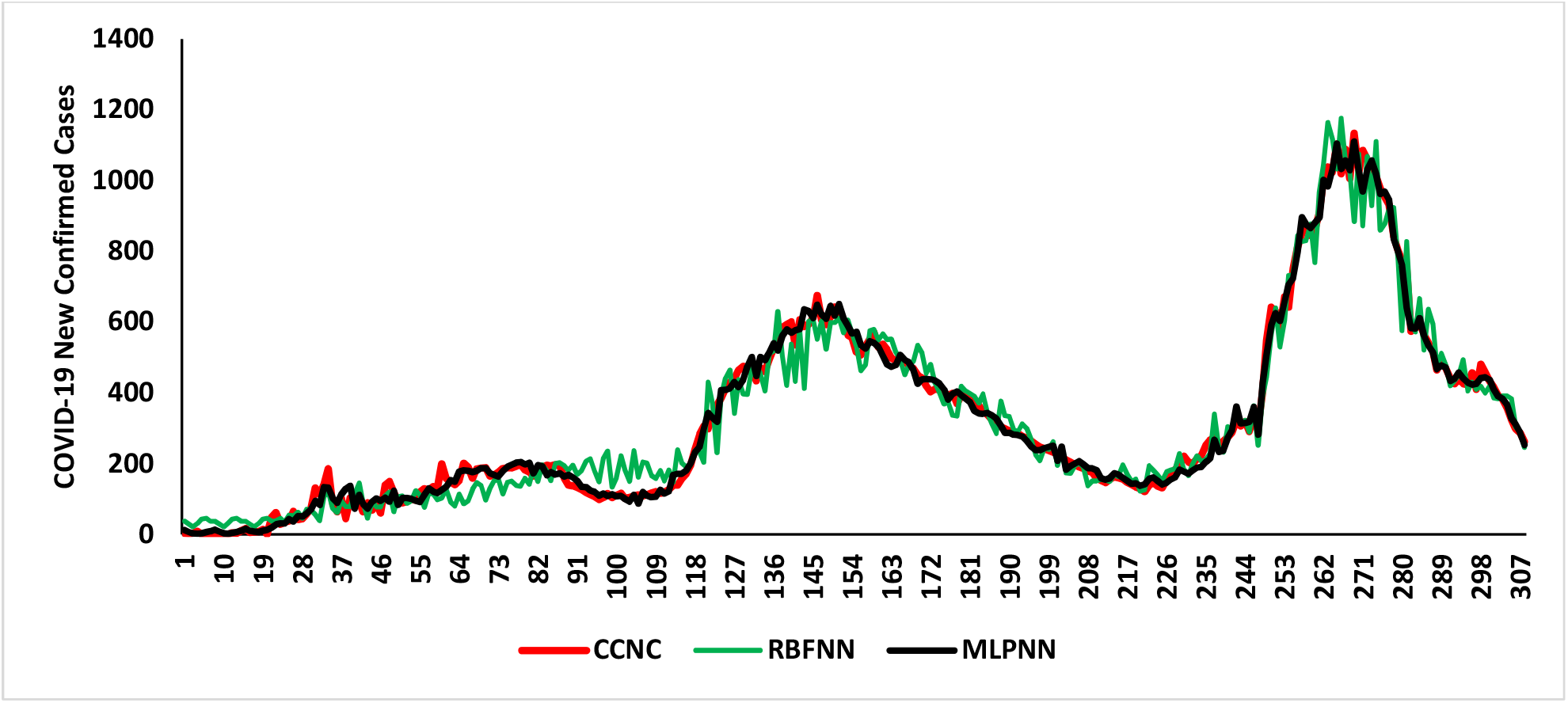
COMPARISON BETWEEN OBSERVED AND PREDICTED VALUES OF CNCC USING RBFNN AND MLPNN MODELS.

To summarize the discussion section, the proposed multi-layer perceptron neural networks was found to have excellent prediction skills for COVIS-19 New Confirmed Cases (CNCC) with regard to the others single artificial neural networks models.

## IV. CONCLUSION

Every day new confirmed cases of COVID-19 are observed in Algeria.

Hence, effective techniques are required to study and analyze the data trends of COVID-19 cases for public health facilities management.

This paper presented the application of different artificial neural networks models for the prediction of COVID-19 New Confirmed Cases (CNCC).

Four different simple neural networks models (GRNN, RBFN, ELM, and MLPNN) were built for CNCC prediction.

The predictive results revealed the potential of MLPNN with a high level of accuracy over the comparable single models for all the considered variables with an exception in terms of performances for ELM models.

The results also depicted that both MLPNN and RBFNN models demonstrated prediction skill and can, therefore, serve as reliable models in public health facilities management.

## Data Availability

The data are available on the official website of the Algeria Health Ministry.

